# Efficacy of the Swede Score in Optimizing Triage of Visual Inspection with Acetic Acid Positive Women

**DOI:** 10.1101/2025.09.20.25336241

**Authors:** Arefa Yesmin, Moshfiqur Rahman, Tannita Das, Afroza Kutubi, Mst. Irin Nahar, Mowmita Zaman Khan, Farhana Mahfuz, Sumona Parvin, Nargis Pervin, Mohammad Azmain Iktidar

## Abstract

**Background:** Conventional screening methods for cervical cancer like visual inspection with acetic acid (VIA) have limited specificity. The Swede score, a structured colposcopic scoring system, may improve detection and triage of cervical intraepithelial neoplasia (CIN) in VIA-positive women.

**Methods:** This cross-sectional study was conducted at the Department of Obstetrics and Gynaecology, Sir Salimullah Medical College, Mitford Hospital, from August 2022 to July 2023. A total of 60 VIA-positive women matching the selection criteria were consecutively enrolled after receiving informed consent. Socio-demographic information and detailed medical history were recorded. All participants underwent colposcopic assessment using the Swede score, followed by colposcopy-guided biopsy. Data were analyzed by Stata (v.17).

**Results:** The mean age was 41.7±8.1 years, with 41.7% aged 40–49 and 48.3% having below-secondary education. Chronic cervicitis was found in 50%, while 30% had CIN1 and 18.3% had CIN2+. The mean Swede score was 5.3±2.0. Notably, 96.6% of women with abnormal histopathology had Swede scores of 5–10, while 61.3% of those with normal histology scored 0–4. A Swede score cut-off of 6 demonstrated high sensitivity (82.8%) and specificity (96.8%) for CIN1+ (PPV 96.0%, NPV 85.7%). For CIN2+, a cut-off of 7 achieved perfect sensitivity (100%) and high specificity (85.7%), with an NPV of 100%. Multivariate analysis showed that each unit increase in Swede score increased the risk of CIN by 39-fold (AOR: 39.14, 95% CI: 2.33–658.33).

**Conclusion:** The Swede score is a reliable tool for detecting CIN among VIA-positive women, supporting its use for targeted biopsies and “see-and-treat” strategies in low-resource settings.

## Introduction

Cervical cancer continues to be a significant global health challenge, ranking as the fourth most common cancer among women worldwide [1]. According to the latest GLOBOCAN 2022 data, there were approximately 662,301 new cases and 348,874 deaths from cervical cancer globally in that year, with substantial geographic disparities highlighting inequalities in access to prevention and treatment [2]. Low- and middle-income countries bear the brunt of this burden, accounting for about 94% of deaths, with the highest incidence and mortality rates observed in sub-Saharan Africa, Central America, and South-East Asia [3, 4]. In Bangladesh, cervical cancer remains a critical issue, contributing to around 12% of female cancers, with historical data showing 8,068 new cases and 5,214 deaths in 2018, though updated figures specific to the country are not yet available in the latest datasets [5].

Over the past few decades, high-income countries have achieved over an 80% reduction in cervical cancer incidence through organized screening programs utilizing tools like colposcopy to detect premalignant lesions [6]. However, such comprehensive systems are often absent in resource-limited settings, where alternative screening methods like Visual Inspection with Acetic Acid (VIA) have become vital due to their affordability and immediacy of results [7]. VIA positive is defined as the presence of fixed, dense, acetowhite patches in the transformation zone near the squamocolumnar junction. Positive cases are referred to higher-level facilities, where colposcopy is performed and colposcopy guided biopsy is taken if needed [8].

Colposcopy, introduced by Hans Hinselmann in 1925, offers a more specific diagnostic approach by providing magnified visualization of the cervix to pinpoint abnormal lesions for biopsy and histopathological confirmation [9]. However, its accuracy can be subjective, depending on the clinician’s experience. To mitigate this, standardized scoring systems like the Reid Colposcopic Index (RCI) and the Swede score have been developed to enhance diagnostic consistency [10, 11]. The Swede score, introduced by Strander et al. in 2005, builds on the RCI by including lesion size alongside parameters such as acetowhiteness, margins, vessel patterns, and iodine staining, yielding a score from 0 to 10. Recent studies confirm its efficacy, with a Swede score of ≥5 showing high sensitivity (up to 100%) for detecting cervical intraepithelial neoplasia (CIN) 1+ lesions, and a score of ≥8 offering exceptional specificity (up to 100%) for high-grade lesions (CIN 2+), supporting a “see-and-treat” approach that minimizes clinic visits and patient drop-out in resource-constrained environments [12].

In Bangladesh, where VIA is a primary screening tool but lacks specificity, and colposcopy access is limited, the Swede score holds significant potential to refine cervical cancer screening protocols. Recent research underscores its predictive value, with a score of ≥5 achieving a sensitivity of 88.4% and specificity of 87.1% for CIN 2+ lesions in some studies, while a score of ≥8 can guide immediate treatment with minimal overtreatment risk [13, 14]. Despite its increasing adoption, local evidence on the Swede score’s correlation with histopathological outcomes in VIA-positive women remains scarce, necessitating further investigation to inform national guidelines.

This study emphasized colposcopic evaluation by the Swede score and histopathological results from colposcopy-guided biopsy of VIA-positive women, which may aid in ensuring proper diagnosis of the pre-invasive stage of carcinoma and help to treat by the “see-and-treat” approach.

## Methods

### Study Design and Setting

This was a cross-sectional, observational study conducted in the Department of Obstetrics and Gynaecology at Sir Salimullah Medical College Mitford Hospital, Dhaka, Bangladesh. The study period was from October 2023 to July 2024.

### Study Population and Sampling

The study targeted women aged 30 to 60 years who presented to the outpatient department for routine cervical cancer screening and were found to be VIA-positive. A purposive (non-probability) sampling technique was used to enroll 60 consecutive VIA-positive women who met the eligibility criteria and consented to participate.

### Inclusion and Exclusion Criteria

Eligible participants were women aged 30–60 years, with a positive VIA test result, no prior treatment for cervical pre-cancer or cancer, and willingness to provide informed written consent. Women were excluded if they had visible cervical growths, a known diagnosis of cervical cancer, were pregnant, experiencing active vaginal bleeding at the time of examination, or if the colposcopic examination was unsatisfactory (defined as inability to visualize the entire squamocolumnar junction). Additional exclusions included severe debilitating disease or a history of pelvic irradiation.

### Data Collection Procedures

Upon enrollment, each participant underwent a structured interview to collect socio-demographic information (age, residence, occupation, education), reproductive and gynecological history (parity, age at first intercourse, contraceptive use, menstrual and obstetric history), and clinical symptoms (e.g., abnormal vaginal discharge, post-coital bleeding). This information was recorded using a pre-tested, standardized questionnaire administered by trained research staff.

### Colposcopic Assessment and Swede Scoring

With proper screening protocols in place, each woman was positioned comfortably in a modified lithotomy position to facilitate optimal visualization of the cervix. After thorough counseling, a medium-sized Cusco’s bivalve self-retaining vaginal speculum was gently inserted; warm, clean water was used as the preferred lubricant, both to warm the metal and to avoid any interference with the interpretation of cervical specimens. Once the speculum was in place and the blades were widely separated, a clear view of the cervix and vaginal fornices was obtained. This maneuver often resulted in eversion of the lips of a multiparous cervix, allowing the lower portion of the endocervical canal to come into view.

Upon exposure of the cervix, the cervical vessel pattern was carefully inspected. The clinician also assessed the nature of cervico-vaginal secretions and documented any obvious findings such as ectropion, polyps, nabothian follicles, transformation zone characteristics, atrophy, inflammation, infection, leukoplakia (hyperkeratosis), condylomata, ulcers, growths, or any other lesions in the vaginal fornices. Three solutions were then sequentially applied to the cervix: normal saline, 3–5% acetic acid, and Lugol’s iodine. Normal saline was first used to remove any obscuring mucus and debris and to moisten the cervix; examination with a green filter enhanced visualization of the cervical angioarchitecture.

Next, a swab soaked in 3–5% acetic acid was applied to the cervix for about one minute. The acetic acid caused coagulation of nuclear proteins, which prevented the transmission of light through dysplastic epithelium, making these areas visible as acetowhite changes. Following this, a Lugol’s iodine-soaked swab was applied to the cervix to assess iodine uptake. Mature squamous epithelium, rich in glycogen, stained dark brown, whereas dysplastic or immature cells, lacking glycogen, appeared yellowish.

All colposcopic findings—including those observed after speculum examination, acetic acid application, green filter assessment, and Lugol’s iodine staining—were meticulously documented and scored using the Swede score system [10]. Each of the five parameters— acetowhiteness, margins and surface, vascular pattern, lesion size, and iodine staining—was assigned a score from 0 to 2, according to established criteria. The sum of these scores yielded a total Swede score ranging from 0 to 10 for each patient. The highest score observed during the examination was recorded. One or more colposcopy-guided punch biopsies were then obtained from areas of abnormal colposcopic appearance, regardless of the Swede score, in all VIA-positive cases suspected of cervical lesions. Hemostasis at the biopsy site was achieved by applying pressure with dry cotton swabs, and Monsel’s paste was used if bleeding was excessive. The biopsy specimens were preserved in formalin and sent to the pathology department of Sir Salimullah Medical College Mitford Hospital for histopathological examination.

### Data Management and Statistical Analysis

All data were checked for completeness and accuracy before entry into a secure, password-protected database. Statistical analyses were performed using Stata (version 17). Descriptive statistics were used to summarize demographic and clinical characteristics. Continuous variables were expressed as mean ± standard deviation (SD), and categorical variables as frequencies and percentages. Associations between categorical variables were assessed using Pearson’s chi-squared test and two independent sample t-test. Receiver Operating Characteristic (ROC) curve analysis was conducted to determine the optimal Swede score cut-off for predicting CIN1+ and CIN2+ lesions. Diagnostic performance was evaluated by calculating sensitivity, specificity, positive predictive value (PPV), and negative predictive value (NPV) for various Swede score thresholds. Statistical significance was set at p < 0.05.

### Ethical Considerations

The study protocol was reviewed and approved by the Institutional Ethics Committee of Sir Salimullah Medical College, Dhaka (Reference No:59.14.1100.031.18.001.23.5120). Written informed consent was obtained from all participants prior to enrollment. All procedures were conducted in accordance with the Declaration of Helsinki and local ethical guidelines. Participant confidentiality was strictly maintained throughout the study, and all data were anonymized prior to analysis.

## Results

The study population primarily comprised women aged 40–49 years (41.7%), with the majority residing in urban areas (76.7%) and working as housewives (86.7%). Educational attainment was generally low, as 78.3% had no formal education or had not completed secondary school. Most participants reported a monthly family income between 20,000 and 50,000 BDT (58.3%). Reproductive history revealed that over half (51.7%) had three or more children, and the mean age at first marriage and first pregnancy were 17.0 and 18.7 years, respectively. A quarter (25%) reported a history of abortion. The majority (95%) were menstruating women, with most reporting a period duration of 3–5 days (47.4%) and a cycle length of 30–35 days (36.8%). Contraceptive use was common (60%), with oral contraceptive pills (43.3%) and barrier methods (30%) being the most frequently used methods (Table 1).

**Table 1:** Characteristics of the study participants (n=60)

| **Variables** | **Frequency** | **Percentage** |
| --- | --- | --- |
| **Age (in years)** |  |  |
| <40 | 23 | 38.3 |
| 40-49 | 25 | 41.7 |
| ≥ 50 | 12 | 20.0 |
| **Residence** |  |  |
| Urban | 46 | 76.7 |
| Rural | 14 | 23.3 |
| **Education** |  |  |
| No formal education | 18 | 30.0 |
| Below Secondary | 29 | 48.3 |
| Secondary or above | 13 | 21.7 |
| **Occupation** |  |  |
| Service holder | 8 | 13.3 |
| Housewife | 52 | 86.7 |
| **Monthly family income (BDT)** |  |  |
| ≤20000 | 21 | 35.0 |
| 20000-50000 | 35 | 58.3 |
| >50000 | 4 | 6.7 |
| **Obstetrical history** |  |  |
| **Number of children** |  |  |
| None | 4 | 6.7 |
| One child | 4 | 6.7 |
| Two children | 21 | 35.0 |
| ≥3 children | 31 | 51.7 |
| **History of abortion** | 15 | 25.0 |
| **Age of first pregnancy, years(mean±SD)** | 18.66±3.50 |  |
| **Menstrual history** |  |  |
| **Age of menarche, years (mean±SD)** | 11.42±1.25 |  |
| **Menstruation** |  |  |
| Post-menopausal women | 3 | 5.0 |
| Menstruating women | 57 | 95.0 |
| **Cycle duration (in days)\*** |  |  |
| 26-28 | 5 | 8.8 |
| 28-30 | 14 | 24.6 |
| 28-32 | 7 | 12.3 |
| 30-35 | 21 | 36.8 |
| 35-40 | 3 | 5.3 |
| Irregular | 7 | 12.3 |
| **Period duration (in days)\*** |  |  |
| 2-3 | 18 | 31.6 |
| 3-5 | 27 | 47.4 |
| 5-7 | 5 | 8.8 |
| 7-10 | 7 | 12.3 |
| **Contraceptive History** |  |  |
| **Use of contraception** |  |  |
| Yes | 36 | 60.0 |
| No | 24 | 40.0 |
| **Method of contraception** |  |  |
| Oral contraceptive pill | 26 | 43.3 |
| Barrier method (condom) | 18 | 30 |
| Subdermal implant | 2 | 3.3 |
| Others | 7 | 11.7 |
| **Clinical History** |  |  |
| Post-menopausal Bleeding | 2 | 3.3 |
| Abnormal Vaginal Discharge | 47 | 78.3 |
| Post-coital Bleeding | 23 | 38.3 |
| Intermenstrual Bleeding | 15 | 25.0 |

\*Only applicable for menstruating women, n=57

The Swede score was significantly associated with histopathological findings. No women having a Swede score of <5 exhibited a lesion of CIN 2 or worse, while all the women having no abnormality or chronic cervicitis had a Swede score of 0-4 (Table 2).

**Table 2:**
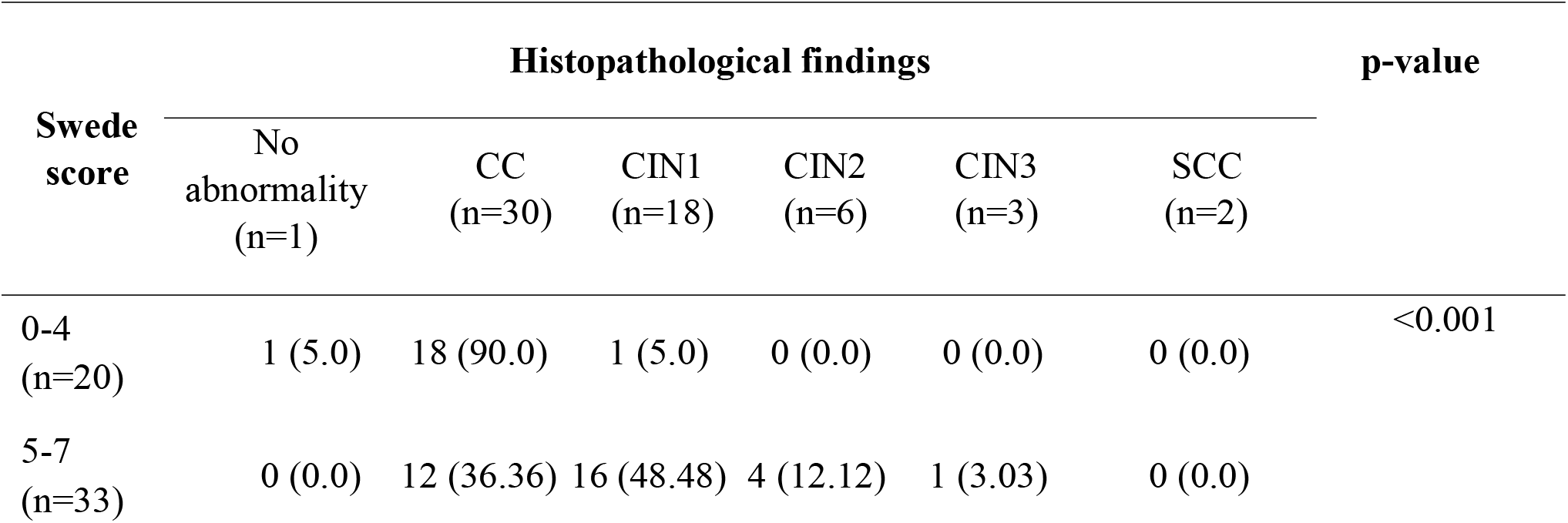
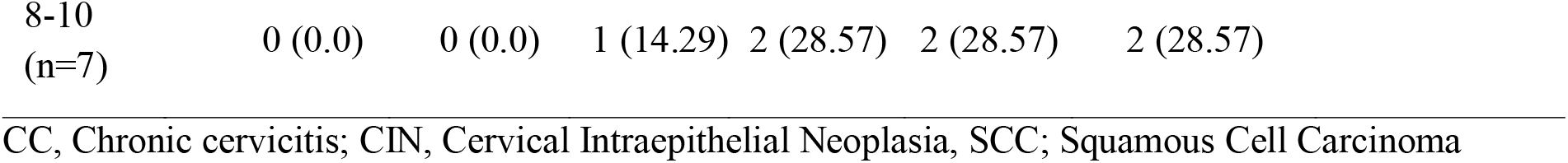
Distribution of Swede score according to histopathological findings of the colposcopy guided biopsy (n=60)

| Swede score | Histopathological findings |  |  |  |  |  | p-value |
| --- | --- | --- | --- | --- | --- | --- | --- |
|  | No abnormality (n=1) | CC (n=30) | CIN1 (n=18) | CIN2 (n=6) | CIN3 (n=3) | SCC (n=2) |  |
| 0-4 (n=20) | 1 (5.0) | 18 (90.0) | 1 (5.0) | 0 (0.0) | 0 (0.0) | 0 (0.0) | <0.001 |
| 5-7 (n=33) | 0 (0.0) | 12 (36.36) | 16 (48.48) | 4 (12.12) | 1 (3.03) | 0 (0.0) |  |

|  |  |  |  |  |  |  |
| --- | --- | --- | --- | --- | --- | --- |
| 8-10<br>(n=7) | 0 (0.0) | 0 (0.0) | 1 (14.29) | 2 (28.57) | 2 (28.57) | 2 (28.57) |

CC, Chronic cervicitis; CIN, Cervical Intraepithelial Neoplasia, SCC; Squamous Cell Carcinoma

Table 3 shows that women with CIN1+ histopathological findings were significantly older and had a longer duration of marriage compared to those with normal or chronic cervicitis (CC) findings. Specifically, the mean age in the CIN1+ group was 44.8 years versus 38.9 years in the normal/CC group (p=0.004), and the mean duration of marriage was 27.3 years versus 22.2 years, respectively (p=0.03).

**Table 3:**
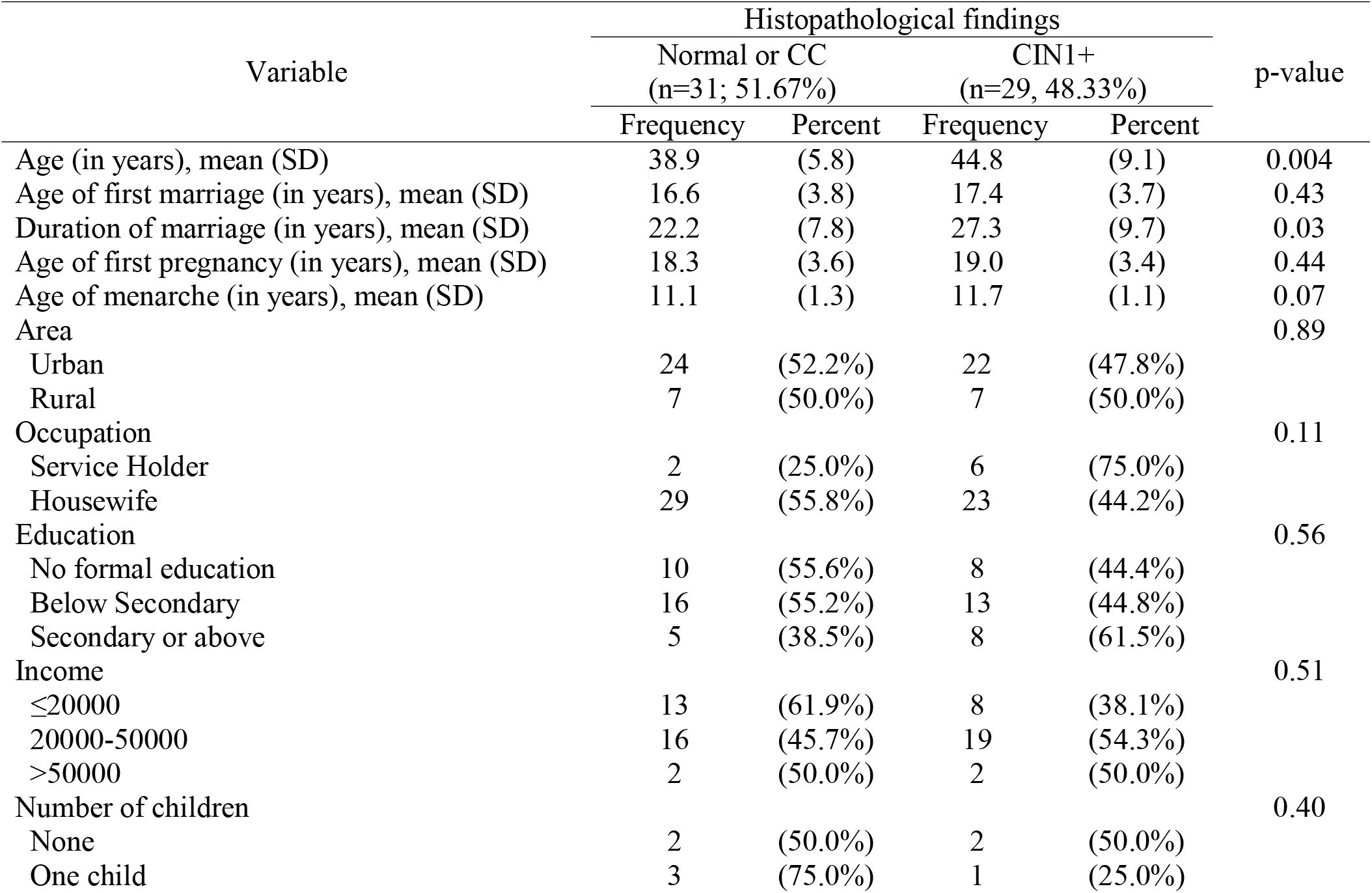
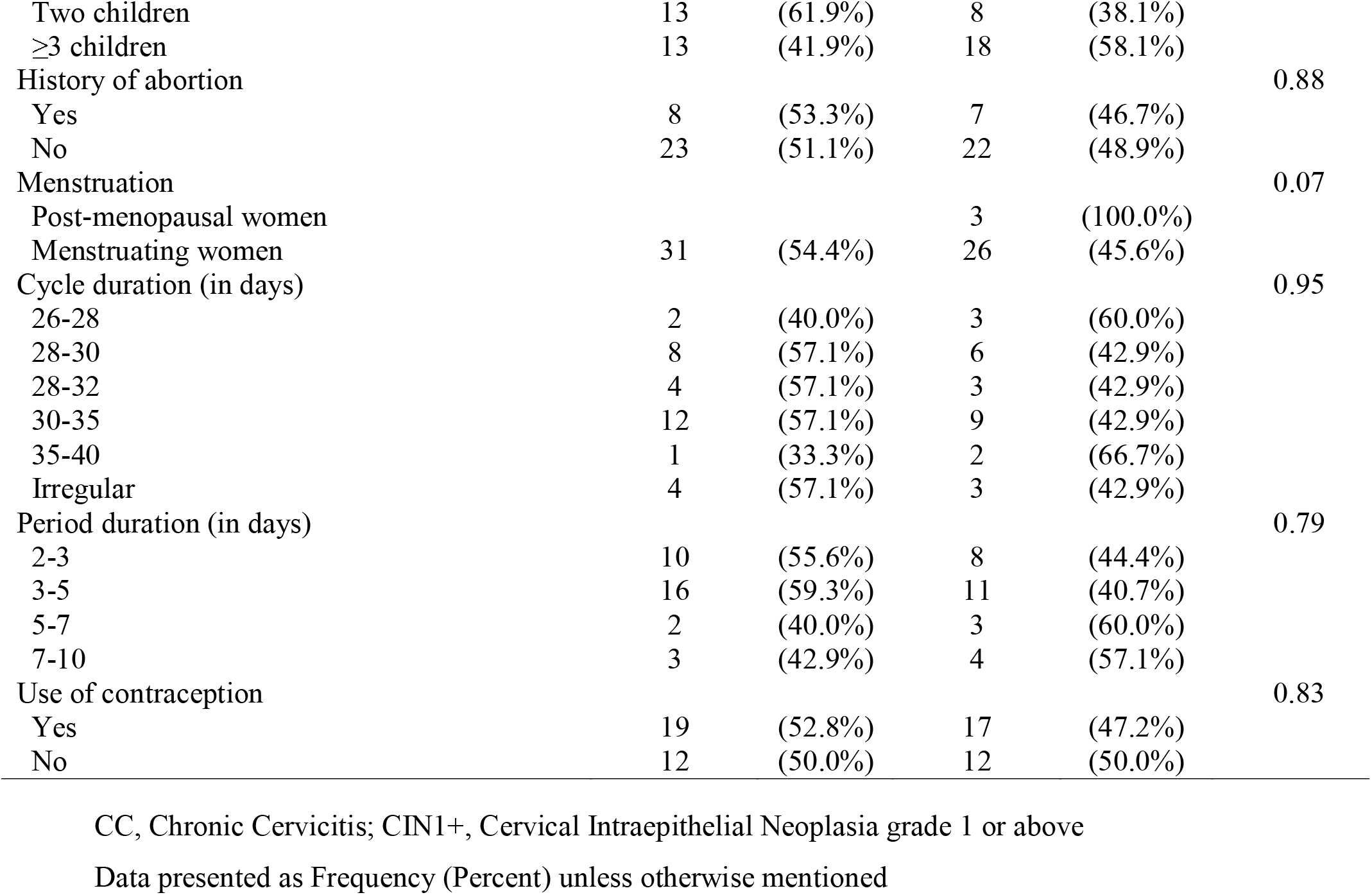
Comparison of Characteristics Between Women With Normal/Chronic Cervicitis and CIN1+ Histopathological Findings (N=60)

Area under the ROC curve (AUC) of Swede score to detect Cervical Intraepithelial Neoplasia grade 1 or above was 0.95 (95%CI: 0.90-0.99) and to detect Cervical Intraepithelial Neoplasia grade 2 or above was 0.96 (95%CI: 0.92-0.99) (Figure 1)

**Figure 1:**
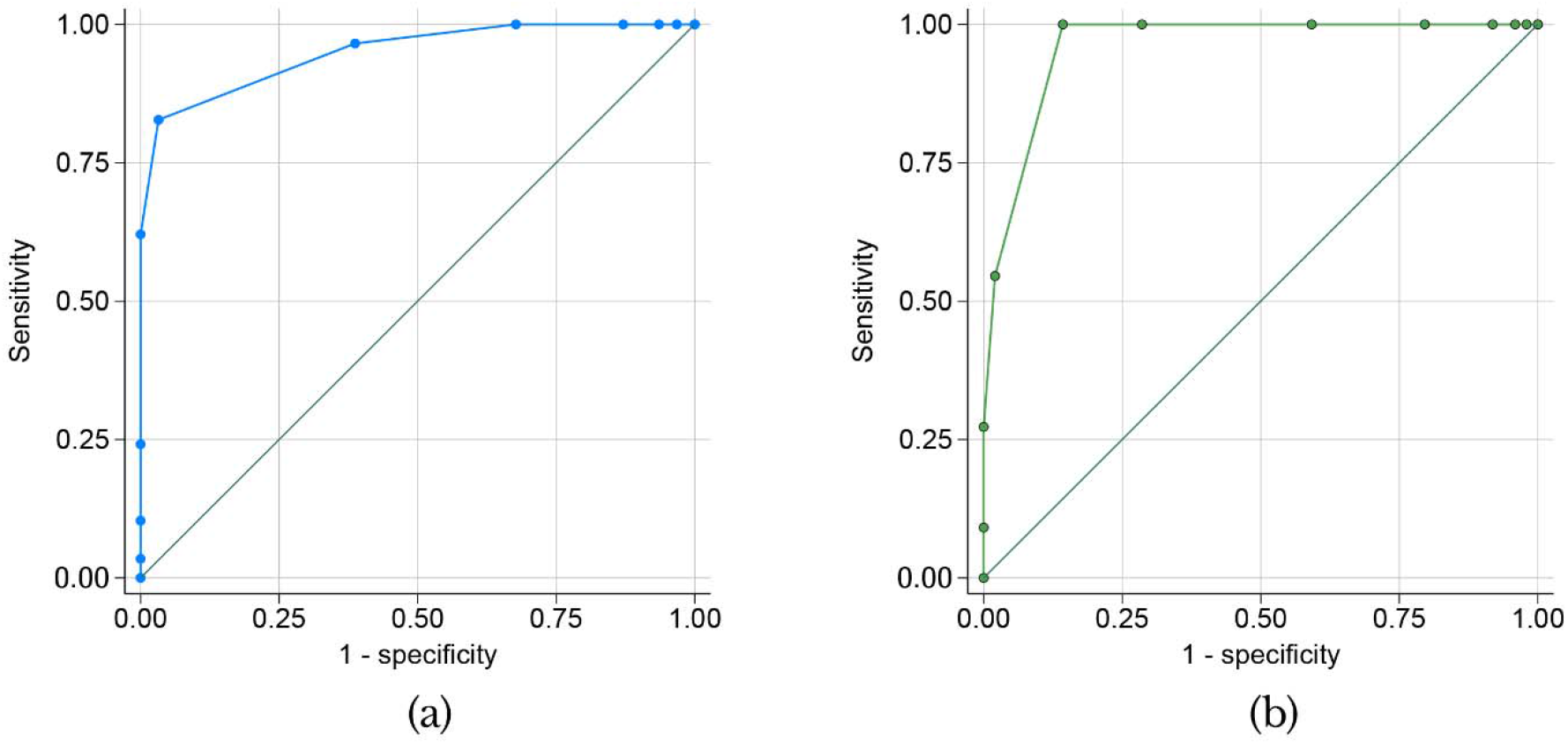
Receiver operator characteristics curve of Swede score to detect (a) Cervical Intraepithelial Neoplasia grade 1 or above (b) Cervical Intraepithelial Neoplasia grade 2 or above.

A Swede score cut-off of 6 showed high sensitivity (82.8%) and excellent specificity (96.8%) for detecting CIN1+, with a strong PPV (96.0%) and NPV (85.7%). For CIN2+, a cut-off of 7 achieved perfect sensitivity (100%) and high specificity (85.7%), with an NPV of 100%, ensuring all true cases were identified and negatives reliably excluded.

Table 4 presents that for each unit increase in Swede Score, the risk of developing CIN increased by 39 times (AOR:39.14, 95% CI: 2.33, 658.33).

**Table 4:**
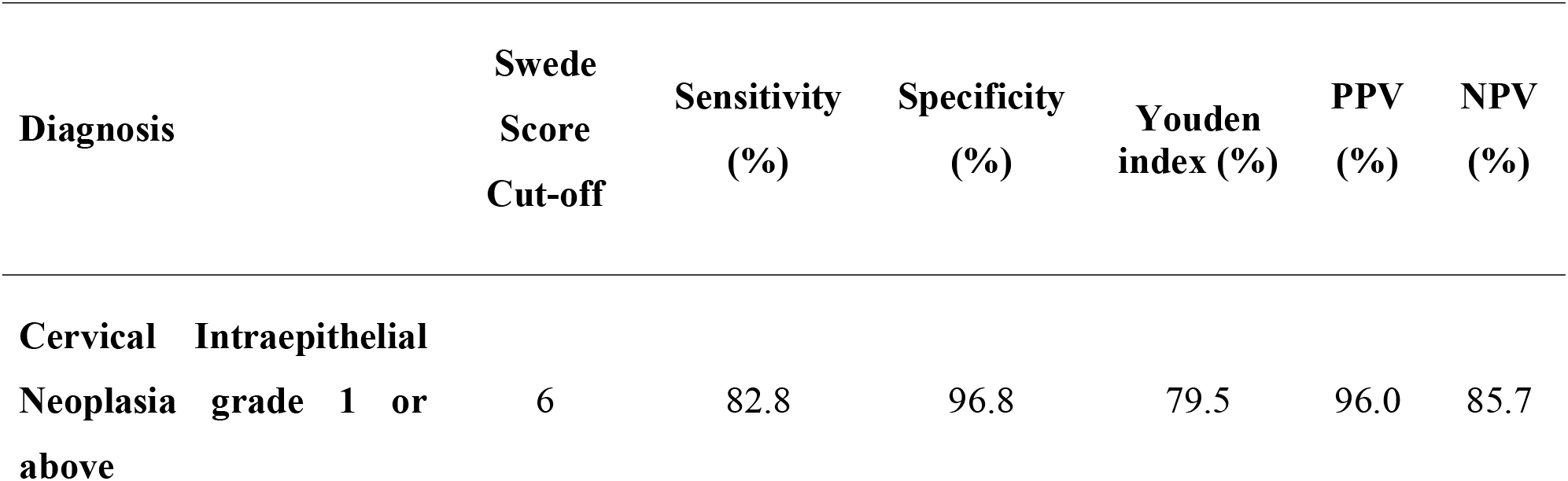
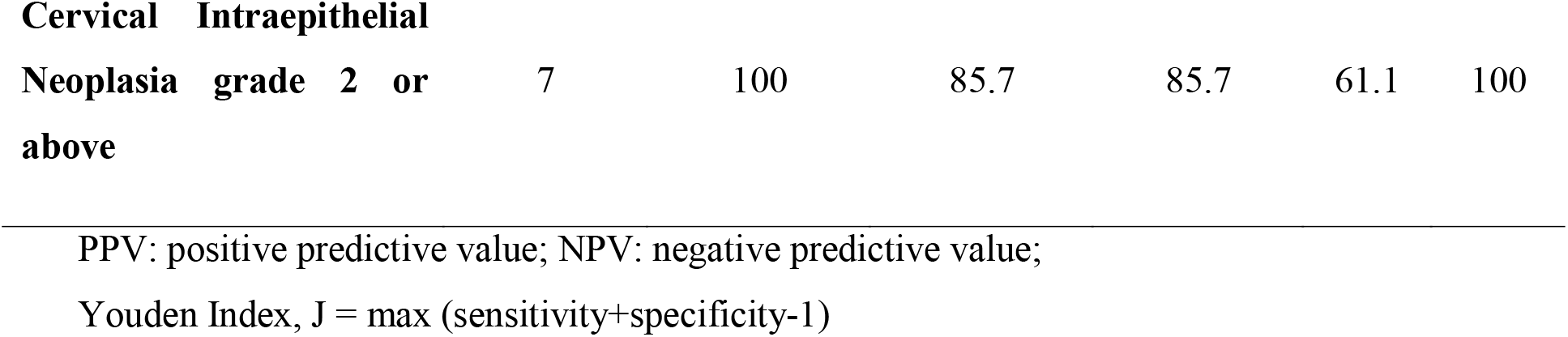
Efficacy of Swede score to detect cervical intraepithelial neoplasia in VIA Positive Women (n=60)

**Table 4:** Multivariate logistic regression results.

| Variable | Adjusted Odds ratio | p-value | 95% Confidence Interval |
| --- | --- | --- | --- |
| Swede Score | 39.14 | 0.011 | 2.33, 658.33 |
| Adjusted for age, age at menarche, age at first pregnancy, number of children, oral contraceptive pill use |  |  |  |

## Discussion

This study assessed the diagnostic utility of the Swede score in identifying cervical intraepithelial neoplasia (CIN) among VIA-positive women in a tertiary care hospital in Bangladesh. The findings demonstrate that the Swede score is a robust and practical tool for triaging women at risk of cervical precancerous lesions, supporting its integration into cervical cancer screening protocols in resource-limited settings.

The mean age of participants in this study was 41.72±8.05 years, with the largest proportion (41.7%) aged 40–49 and nearly half (48.3%) having less than secondary education. These demographics are similar to those found by Agarwal et al. and Rahman et al., who also observed that VIA positivity is linked to older age and lower educational levels [13, 15]. Most women presented with abnormal vaginal discharge (78.3%), followed by post-coital bleeding (38.3%), moderate dyspareunia, intermenstrual bleeding (25%), and postmenopausal bleeding (3.3%), consistent with findings from other Bangladeshi and regional studies [16].

The present study reveals that women with CIN1+ histopathological findings were significantly older (mean age 44.8 vs. 38.9 years) and had a longer duration of marriage (27.3 vs. 22.2 years) compared to those with normal or chronic cervicitis (CC) findings, with both differences reaching statistical significance (p=0.004 and p=0.03, respectively). These demographic associations suggest that cumulative exposure to risk factors, such as persistent HPV infection, increases with age and marital duration, a finding similar to Agarwal et al., who also reported higher rates of CIN among older, multiparous women with lower educational status [17, 18]. This underlines the need for targeted screening in populations with similar sociodemographic profiles.

Chronic cervicitis was diagnosed in half of the women, while 48.3% had abnormal histopathology, mainly CIN1 (30%) and CIN2+ (18.3%). Similar proportions were reported in Indian studies, where chronic cervicitis ranged from 39.3% to 65.45%, CIN1 from 21.4% to 27.5%, and CIN2+ from 10.9% to 25% [12]. The mean Swede score in this cohort was 5.28±1.98. Nearly all women with abnormal histopathology had Swede scores between 5 and 10 (96.6%), while 61.3% of those with normal histopathology scored 0–4. No participant with a Swede score below 6 had CIN2 or worse, and all women with normal or chronic cervicitis had scores between 0 and 6. This pattern is consistent with findings from Agarwal et al., Rahman et al., and Penumalli et al., who reported that Swede scores of 0–4 were typically associated with normal histology, while higher scores correlated with more severe lesions [13, 15, 19].

A significant association was observed between Swede score and histopathological grade (p < 0.001), with women having abnormal histopathology showing significantly higher Swede scores. Previous research has similarly validated the Swede score as a reliable marker for lesion severity, with studies by Ding et al. and Penumalli et al. demonstrating strong agreement between Swede score-based colposcopic impressions and histopathology [19, 20].

A Swede score cut-off of ≥6 was optimal for predicting abnormal histopathology (CIN1+), yielding sensitivity, specificity, PPV, NPV, and accuracy of 82.8%, 96.8%, 79.5%, 96.0%, and 85.7, respectively. These values are comparable to other studies, such as those by Rahman et al., Karya et al., and Ranga et al., which reported high sensitivity and negative predictive value for this threshold [13, 14, 21]. For high-grade lesions (CIN2+), a Swede score of ≥7 achieved 100% sensitivity, 85.71% specificity, 61.11% PPV, and 100% NPV. Agarwal et al. also found that a Swede score of 8 or more had 100% specificity for high-grade lesions, though with a trade-off in sensitivity and overall accuracy depending on the cut-off used [15]. In this study, a Swede score of ≥8 had 54.55% sensitivity, 97.96% specificity, 85.71% PPV, 90.57% NPV, and 90% accuracy for detecting CIN2+, supporting the use of higher cut-offs for direct treatment without preoperative biopsy and minimizing overtreatment. Overall, the Swede score enables more targeted biopsies and supports a “see-and-treat” approach, improving case detection and reducing unnecessary procedures.

However, there is ongoing debate regarding the ideal balance between sensitivity and specificity, especially in low-resource settings. Chowdhury et al noted that raising the Swede score cut-off increases specificity but at the expense of sensitivity, potentially missing a subset of high-grade lesions [18]. In our cohort, a cut-off of 7 or higher offered perfect sensitivity and high specificity, but the positive predictive value, while strong, was not absolute. This trade-off highlights the importance of local validation and the need to tailor cut-offs to the clinical context and patient population.

The Swede score’s simplicity and reproducibility are additional strengths, as emphasized by recent reviews [22]. Unlike the Reid Colposcopic Index, the Swede score incorporates lesion size, which has been shown to correlate strongly with histological severity and improves the score’s predictive accuracy [23]. Comparative studies have found that both scores perform well, but the Swede score’s structure may make it more accessible for clinicians with varying levels of colposcopic experience [12].

Importantly, the present study also confirms the Swede score’s utility in reducing unnecessary biopsies and supporting immediate treatment decisions. With a cut-off of 8, specificity for CIN2+ approached 100% in several studies, suggesting that excisional treatment can be safely offered at this threshold, minimizing overtreatment and loss to follow-up [24]. This is particularly relevant in Bangladesh, where repeated clinic visits can be a barrier to care.

Despite these strengths, limitations remain. The study’s single-center design and moderate sample size may affect generalizability. Inter-observer variability, a known challenge in colposcopy, was not formally assessed, though the Swede score’s structured criteria may help mitigate this.

## Conclusion

In conclusion, this study supports the Swede score as a reliable, evidence-based tool for guiding the management of VIA-positive women. Its high sensitivity and specificity at appropriate cut-offs, ease of use, and potential to streamline “see-and-treat” protocols make it a valuable addition to cervical cancer prevention strategies in low- and middle-income countries

## Data Availability

All data produced in the present study are available upon reasonable request to the authors.

## Statements & Declarations

### Author Contributions

All authors contributed to the study conception and design. Material preparation, data collection, and analysis were performed by Moshfiqur Rahman, Tannita Das, Afroza Kutubi, Mst. Irin Nahar, Mowmita Zaman Khan, Farhana Mahfuz, and Sumona Parvin. The first draft of the manuscript was written by Arefa Yesmin, Nargis Pervin and Mohammad Azmain Iktidar, and all authors commented on previous versions of the manuscript. All authors read and approved the final manuscript.

### Ethics approval

This study was performed in line with the principles of the Declaration of Helsinki. Approval was granted by the Institutional Ethics Committee of Sir Salimullah Medical College, Dhaka (Reference No:59.14.1100.031.18.001.23.5120).

### Consent to participate

Informed consent was obtained from all individual participants included in the study.

### Consent to publish

Not applicable.

